# Investigating disorder-specific and transdiagnostic alterations in model-based and model-free decision-making

**DOI:** 10.1101/2023.12.11.23299814

**Authors:** Franziska Knolle, Pritha Sen, Adam Culbreth, Kathrin Koch, Benita Schmitz-Koep, Deniz A. Gürsel, Klaus Wunderlich, Mihai Avram, Götz Berberich, Christian Sorg, Felix Brandl

**Affiliations:** School of Medicine and Health, Department of Diagnostic and Interventional Neuroradiology, Technical University of Munich, Munich, Germany; School of Medicine and Health, TUM-NIC Neuroimaging Center, Technical University of Munich, Munich, Germany; Department of Psychiatry, University of Cambridge, Cambridge, UK; Department of Psychiatry, Maryland Psychiatric Research Center, University of Maryland, School of Medicine, Baltimore, USA; Translational Psychiatry, Department of Psychiatry and Psychotherapy, University of Lübeck, Lübeck, Germany; Windach Institute and Hospital of Neurobehavioural Research and Therapy, Windach, Germany; Technical University of Munich, School of Medicine, Department of Psychiatry, Munich, Germany

**Author notes:** corresponding author: Dr. Franziska Knolle, Department of Diagnostic and Interventional Neuroradiology, School of Medicine and Health, Klinikum rechts der Isar, Technical University Munich, 81675 Munich, Germany.

**Keywords:** Transdiagnostic, Schizophrenia, Depression, Obsessive-compulsive disorder, Model-based, Model-free, Win-stay, Decision-Making, Reward Processing, Two-step task

## Abstract

**Background:** Decision-making alterations are present in psychiatric illnesses like major depressive disorder (MDD), obsessive-compulsive disorder (OCD), and schizophrenia, linked to symptoms of the respective disorders. Understanding unique and shared decision-making alterations across these disorders is crucial for early diagnosis and treatment, especially given potential comorbidities.

**Methods:** Using two computational modeling approaches – logistic regression and hierarchical Bayesian modeling – we analyzed alterations in model-based and model-free decision-making in a transdiagnostic cohort comprising MDD (N=23), OCD (N=25), and schizophrenia (N=27) patients. Our aim was to identify disorder-specific and shared alterations and their associations with symptoms.

**Results:** Overall, participants of all groups relied on model-free decision-making. Our results revealed that schizophrenia patients had the lowest learning rate and highest switching rate, indicating low perseverance. Further, OCD patients were more random in both task stages compared to controls and MDD patients. All patient groups exhibited more randomness in responses than controls, with schizophrenia patients showing the highest levels. Importantly, the study showed that increased model-free behavior correlated with elevated depressive symptoms and more model-based decision making was linked to lower anhedonia levels across all patients.

**Conclusions:** This study highlights disorder-specific and shared decision-making alterations in individuals with MDD, OCD, and schizophrenia. This study suggests that anhedonia and depressive symptoms, which are present in all three disorders, share underlying behavioral mechanisms. Improving model-based behavior may which may be a target for intervention and treatment.

## Introduction

Decision-making and learning impairments are core characteristics of a wide range of psychiatric disorders^1–3^ such as major depressive disorder (MDD)^4,5^, obsessive-compulsive disorder (OCD)^6,7^, and schizophrenia^8,9^ and link to their specific symptoms. As these disorders share symptoms, it is important to identify decision-making alterations which are specific to each disorder and those that occur across several disorders. A better understanding of these associations may facilitate therapeutic rehabilitation strategies for these patients.

MDD is characterized with persistent symptoms like anhedonia, rumination, and cognitive biases^10–12^. Decision-theoretic approachs^13,14^ suggest that alterations in reward anticipation^15,16^, effort-cost evaluation^17–19^, and a negative bias^20,21^ could be underlying mechanisms. A study by Ang and colleagues^22^ revealed that MDD patients show steeper discounting of effort-based rewards and reduced willingness to invest cognitive effort. Similarly, Treadway and colleagues^19^ demonstrated that MDD patients were less inclined to expend efforts for rewards and less able to use information about reward magnitude and probabilities for optimal decision-making, and that motivational deficits were associated with higher symptom severity. Furthermore, general deficits in reinforcement learning especially the brain signals in response to reward prediction error and expected value (i.e., wanting), decreased reward sensitivity (i.e., liking) and model-free (i.e., habitual) or model-based (i.e., goal-directed) learning have been proposed^23–26^. In probabilistic reward tasks, MDD patients lack the typical preference for frequently rewarded choices, which correlates with anhedonia scores^27,28^. These findings highlight the involvement of reward-related mechanisms in core MDD symptoms, as revealed by cognitive neuroscience approaches.

OCD is characterized by chronic doubting, and compulsive behaviors reflected in dysfunctional decision-making^29^. OCD patients exhibit difficulty in adapting reward perception and tend to make rigid, repetitive decisions^30^. Some studies suggesting that OCD may result from dysfunctional goal-directed control and an over-reliance on habitual control^6,31,32^. The literature presents conflicting evidence regarding the associations between symptom severity and decision-making abnormalities^6,33–36^. This inconsistency may be partially explained by effects of illness duration^36^. In a probabilistic learning paradigm, Murray and colleagues^37^ reported that dysfunctions during reward processing in OCD may be of dopaminergic origin, which may be similar to those alterations seen in schizophrenia patients^38–40^. Furthermore, in a two-step decision-making task, OCD individuals exhibited more model-free choices in rewarded outcomes and more model-based choices in loss outcomes, with compulsions severity linked to habitual learning of rewarded outcomes and obsessions severity linked to increased choice switching following losses^41^. Various imaging studies have also indicated altered activations in brain regions that are associated with habit formation^42–45^, highlighting the importance of investigating decision-making alterations in OCD.

In schizophrenia patients, dysfunctional decision-making relates to both positive (hallucinations and delusions) and negative symptoms (apathy, social withdrawal)^8,46^. Delusions have been associated with “jumping-to-conclusions,” which may lead to hasty judgments and false beliefs^47–49^. Deficits in reward anticipation^50–52^, prediction error processing^38–40^ and reward and punishment sensitivity^53–55^ have been shown in early and late stages of schizophrenia. Furthermore, alterations reward processing have been found to contribute to diminished motivation, affecting future reward assessment^56–58^. Impaired habitual decision-making has also been identified as a contributing factor to negative symptoms in schizophrenia^59^. Impaired effort-based decision-making may be linked to motivational anhedonia in psychosis as patients are unwilling to expend effort to gain rewards^60–62^. Compared to controls, schizophrenia patients exhibit reduced reliance on goal-directed decision-making, indicating more unpredictable behavior^63^.

The literature suggests alterations in distinct reinforcement learning systems in psychiatric disorders like OCD, MDD, and schizophrenia^59,63–68^, which underlie model-free and model-based decision-making processes^69^. These processes can be studied using the two-step Markov decision task^69^, yet no study has examined these in a transdiagnostic sample. Our study aims to identify disorder-specific and general decision-making alterations in MDD, OCD, and schizophrenia patients in remission, using computational modeling and linking them to symptoms. We hypothesized MDD patients to perform better with a mix of model-based and model-free strategies, OCD patients to exhibit more model-free decisions, and schizophrenia patients to be more random in their choices compared to the other patient groups, respectively. Decision-making impairments were expected to correlate with symptoms within each group, with co-occurring symptoms showing similar trends. For instance, depression scores in MDD and schizophrenia patients were expected to correlate similarly with decision-making parameters.

## Methods

### Participants

Twenty-five healthy controls (HC), 23 MDD patients, 25 OCD patients, and 27 schizophrenia (SCZ) patients in psychotic remission took part in the study. Sample sizes were determined based on a power analysis (G*Power 3.1 software^70^) with alpha=0.05, power=0.85, medium effect size of f=0.38 which revealed 90 participants. To account for poor data or outliers, we collected ∼15% more participants per group. The demographic characteristics, cognitive and clinical scores are summarized in Table 1. Schizophrenia and MDD patients were recruited from the Clinic and Polyclinic for Psychiatry and Psychotherapy in Klinikum Rechts der Isar at the Technical University of Munich (TUM), and the OCD patients were recruited from the Windach Institute and Hospital of Neurobehavioral Research and Therapy. Patients met Diagnostic and Statistical Manual of Mental Disorders (DSM)-IV criteria for SCZ, MDD, and OCD^71^. For schizophrenia patients, psychotic remission was based on Andreasen criteria^72^, with a score of ≤3 for each Positive and Negative Syndrome Scale (PANSS) positive item. MDD patients were in an acute depressive episode. Medication for all patients was kept stable for at least two weeks before assessments. Except for Obsessive-Compulsive Inventory – Revised^73^ (OCI-R) scores which was collected only for OCD patients, all other clinical scores were collected across all participants. Healthy controls were age and gender matched and did not report any personal history of severe neurological, psychiatric or medical disorders. The study was approved by the TUM ethics committee. All participants gave their informed, written consent after receiving a complete description of the study.

**Table 1.**
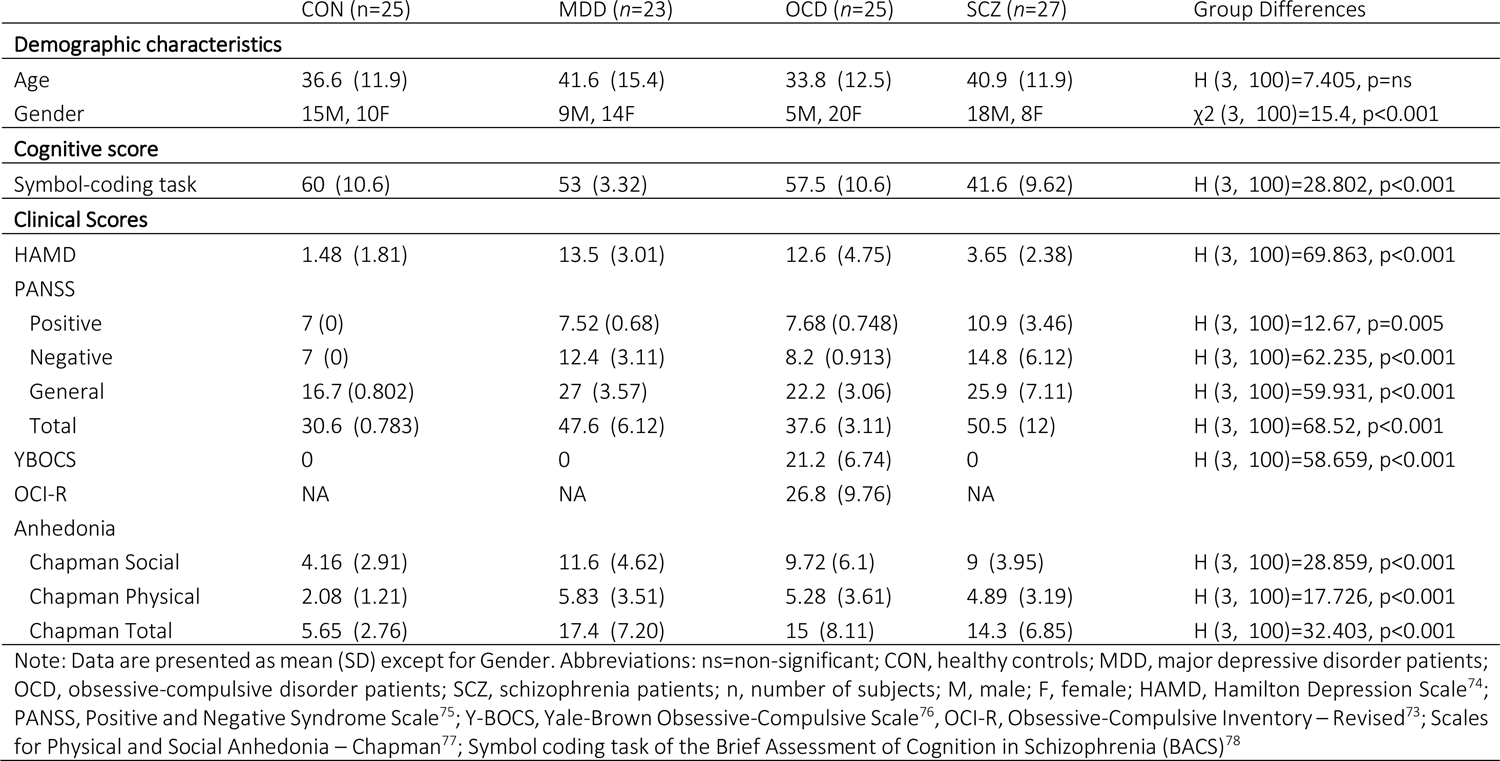
Demographic characteristics, clinical characteristics, and the cognitive function of the healthy controls and patients with major depressive disorder, obsessive-compulsive disorder, and schizophrenia.

|  | CON (n=25) | MDD (n=23) | OCD (n=25) | SCZ (n=27) | Group Differences |
| --- | --- | --- | --- | --- | --- |
| **Demographic characteristics** |  |  |  |  |  |
| Age | 36.6 (11.9) | 41.6 (15.4) | 33.8 (12.5) | 40.9 (11.9) | H (3, 100)=7.405, p=ns |
| Gender | 15M, 10F | 9M, 14F | 5M, 20F | 18M, 8F | $\chi^2$ (3, 100)=15.4, p<0.001 |
| **Cognitive score** |  |  |  |  |  |
| Symbol-coding task | 60 (10.6) | 53 (3.32) | 57.5 (10.6) | 41.6 (9.62) | H (3, 100)=28.802, p<0.001 |
| **Clinical Scores** |  |  |  |  |  |
| HAMD | 1.48 (1.81) | 13.5 (3.01) | 12.6 (4.75) | 3.65 (2.38) | H (3, 100)=69.863, p<0.001 |
| PANSS |  |  |  |  |  |
| Positive | 7 (0) | 7.52 (0.68) | 7.68 (0.748) | 10.9 (3.46) | H (3, 100)=12.67, p=0.005 |
| Negative | 7 (0) | 12.4 (3.11) | 8.2 (0.913) | 14.8 (6.12) | H (3, 100)=62.235, p<0.001 |
| General | 16.7 (0.802) | 27 (3.57) | 22.2 (3.06) | 25.9 (7.11) | H (3, 100)=59.931, p<0.001 |
| Total | 30.6 (0.783) | 47.6 (6.12) | 37.6 (3.11) | 50.5 (12) | H (3, 100)=68.52, p<0.001 |
| YBOCS | 0 | 0 | 21.2 (6.74) | 0 | H (3, 100)=58.659, p<0.001 |
| OCI-R | NA | NA | 26.8 (9.76) | NA |  |
| Anhedonia |  |  |  |  |  |
| Chapman Social | 4.16 (2.91) | 11.6 (4.62) | 9.72 (6.1) | 9 (3.95) | H (3, 100)=28.859, p<0.001 |
| Chapman Physical | 2.08 (1.21) | 5.83 (3.51) | 5.28 (3.61) | 4.89 (3.19) | H (3, 100)=17.726, p<0.001 |
| Chapman Total | 5.65 (2.76) | 17.4 (7.20) | 15 (8.11) | 14.3 (6.85) | H (3, 100)=32.403, p<0.001 |

Note: Data are presented as mean (SD) except for Gender. Abbreviations: ns=non-significant; CON, healthy controls; MDD, major depressive disorder patients; OCD, obsessive-compulsive disorder patients; SCZ, schizophrenia patients; n, number of subjects; M, male; F, female; HAMD, Hamilton Depression Scale<sup>74</sup>; PANSS, Positive and Negative Syndrome Scale<sup>75</sup>; Y-BOCS, Yale-Brown Obsessive-Compulsive Scale<sup>76</sup>; OCI-R, Obsessive-Compulsive Inventory – Revised<sup>73</sup>; Scales for Physical and Social Anhedonia – Chapman<sup>77</sup>; Symbol coding task of the Brief Assessment of Cognition in Schizophrenia (BACS)<sup>78</sup>

### Task description

All participants completed the two-step task adapted from Daw et al^69^ (Figure 1), designed to differentiate between model-free and model-based decision-making (see detailed description in supplements). The task involved maximizing monetary reward over 200 trials, split into four blocks of 50 trials each. Each trial consisted of two decision stages, with participants choosing between two fractal images. Transition probabilities between stages were probabilistic, with common transitions occurring 70% of the time and uncommon transitions 30% of the time. Participants developed strategies based on feedback in stage two to maximize reward. Model-based decision-making involved using transition probabilities to infer action likelihood, while model-free decision-making relied solely on trial outcomes. For detailed imaging results, refer to ^59^, Brandl et al.(in submission), Sen et al.(in submission).

**Figure 1.**
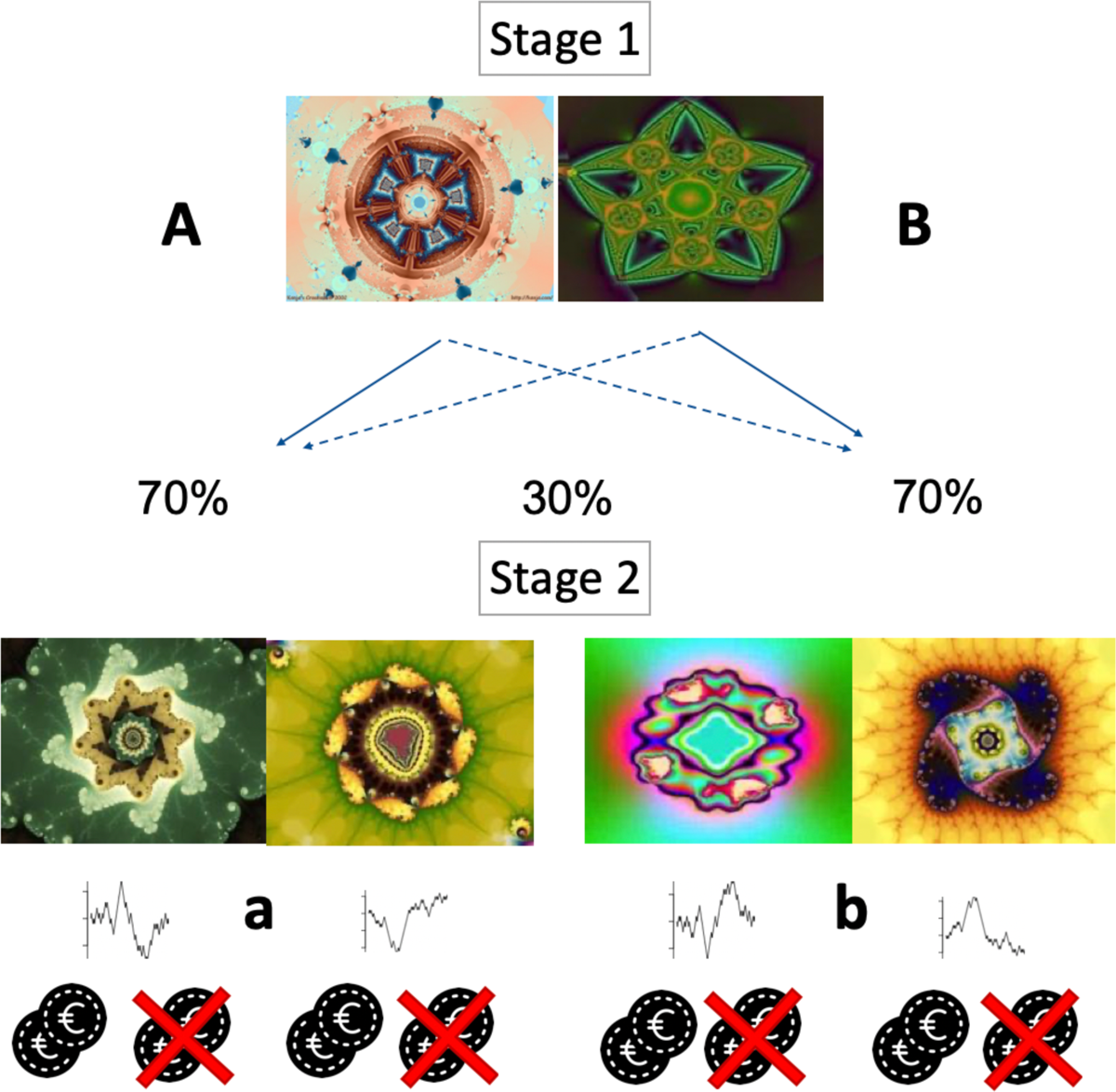
Two-step Markov decision task. Showing the state transition structure which allows discrimination between model-based and model-free behavior. Note: All stimuli in stage 2 are associated with a probabilistic reward changing slowly and independently based on Gaussian random walks, forcing subjects to continuously learn and explore the second stage choices.

### Analysis

#### Stay Probability

Stay-probability was assessed to investigate an action bias. It is the probability of an individual selecting the same first stage choice as in the previous trial, which can show a model-free or a model-based strategy depending on the results of the previous trial (i.e., outcome and transition type). See supplements for exact description. Stay-probabilities were compared across groups using a robust mixed ANOVA with stay probability as the dependent variable, group as the between-subject variable, and previous trial transition type (common/uncommon) and previous trial outcome (rewarded/unrewarded) as within-subject variables. For post-hoc analysis for the factors that resulted in a significant effect, we performed pairwise t-tests corrected for multiple comparisons using Bonferroni adjustments for each of the parameters.

#### Logistic Regression Analysis

We used a hierarchical logistic regression to estimate in a classical approach model-free and model-based decision-making by including independent contributions of choices (stay/shift) on the subject level depending on outcome (rewarded/unrewarded) and transition type of the previous trial (common/ uncommon), similar to previous reports^63,79^. We controlled for BACS and illness duration as fixed effects. We used the following equation:

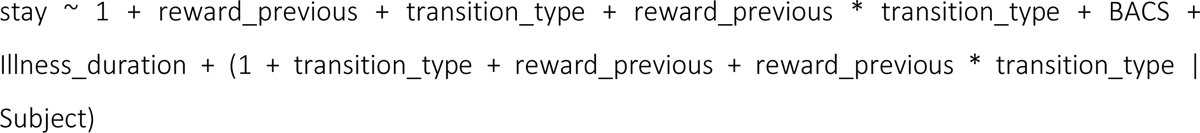

We extracted individual beta estimates for reward reflecting model-free behavior and for the interaction of reward and transition reflecting model-based behavior for each individual subject to explore which decision-making strategy individuals were more likely to use.

We performed a robust mixed ANOVA and robust post hoc tests for significant effects based on trimmed means using the bootstrap method, with beta estimates as the dependent variable, group as the between-subject factor and decision-making strategy (reward beta=model-free/interaction beta= model-based) as the within-subject factor. We then conducted post-hoc analyses for significant effects using one-way robust ANOVA with the bootstrap method.

To explore associations of the beta estimates with clinical scores, we computed, across and within all groups, Spearman’s correlations. We excluded one participant from the MDD group for this analysis due to missing values.

#### Computational Modeling

Furthermore, we used hierarchical Bayesian modeling to estimate computational parameters describing neurocognitive processes underlying decision-making behavior. We fitted four models under one hierarchical prior across all subjects independent of group using the hBayesDM package (version 1.2.1)^80^. We fitted four chains for each model with 1,000 burn-in samples and 3,000 samples. All models are shown in Table 2. The winning model – the 6-parameter model – was determined based on model-convergence with Rhat < 1.04, pareto k < 0.7 and lowest leave one out information criterion (LOOIC) values.

**Table 2.**
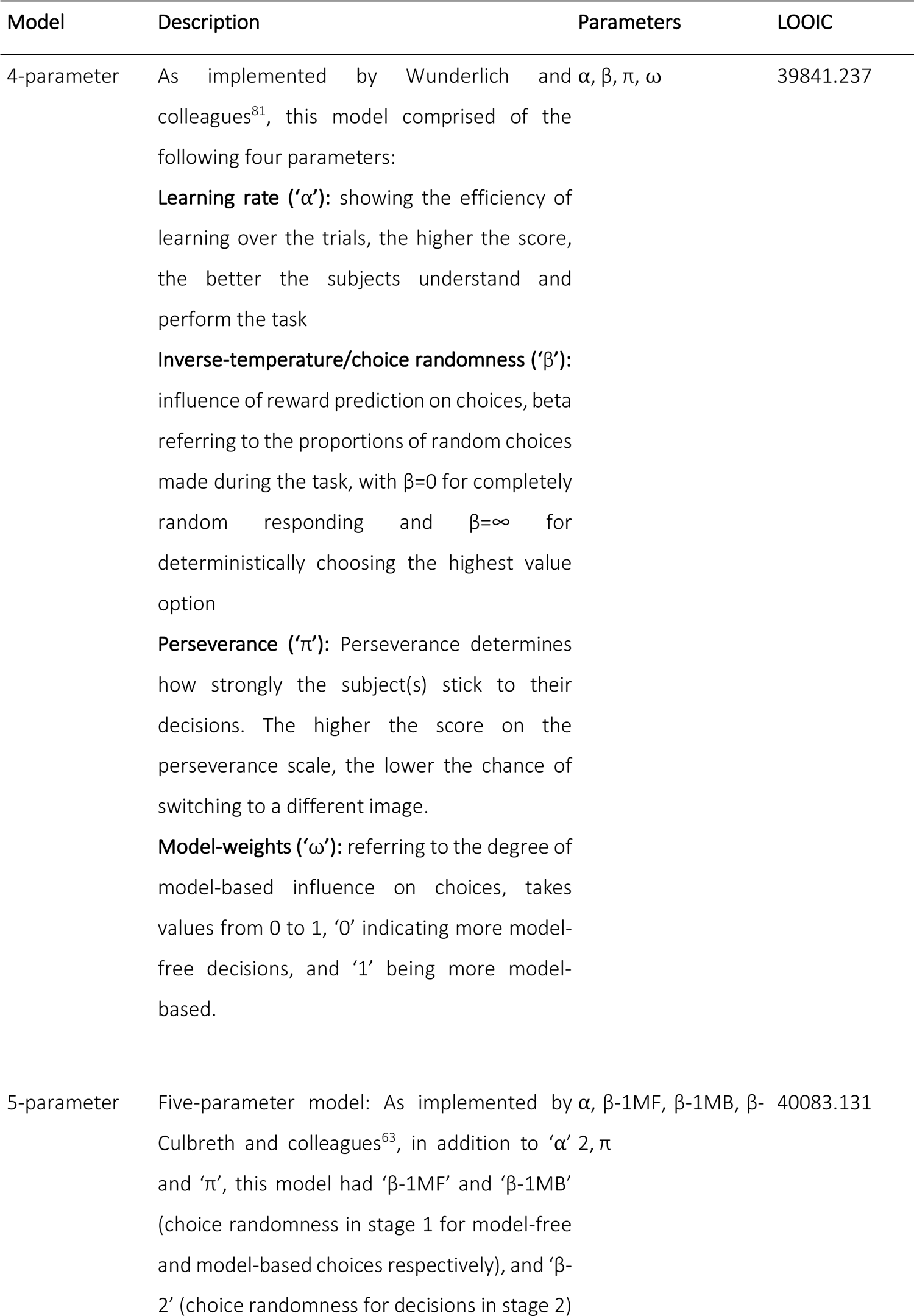
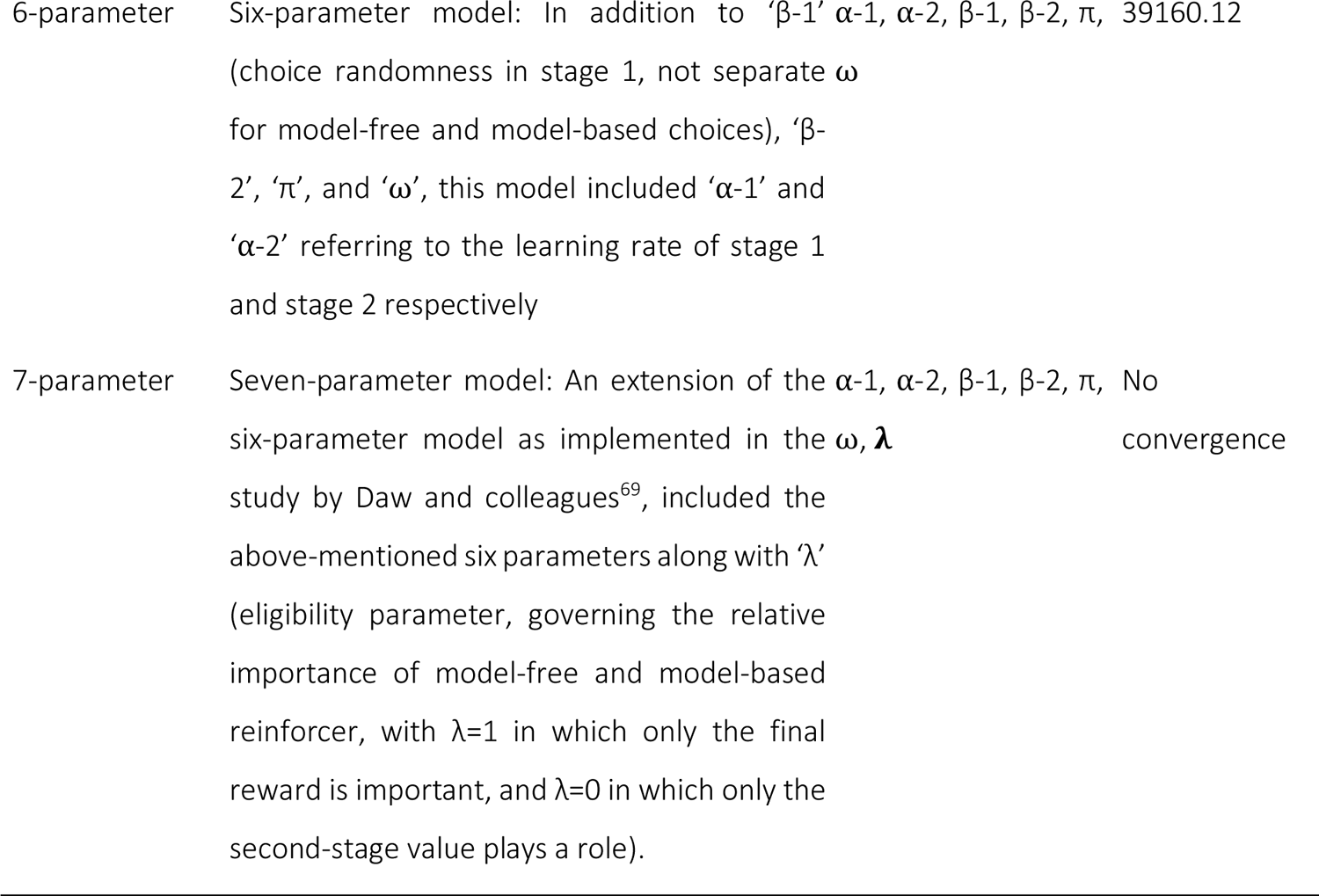
Models with LOOIC values.

To study the associations between modeling parameters and clinical scores, we performed partial Spearman’s correlations controlling for cognitive function using the symbol-coding task of the BACS, as decision-making is highly dependent on cognitive abilities, and illness duration. These correlations were performed across and within all groups. We focused on the parameters model weights ⍵ to reflect the concept of model-free (<0.5) and model-based (>0.5) behavior more closely, and on perseverance π which reflects stickiness or the ability of being able to change a response pattern. We had to exclude one participant from the MDD group for this analysis due to missing values. See supplements for additional correlations.

#### General procedure

All statistical analyses were conducted using the R Statistical Software (version 4.1.1)^82^. Data was visualized using the ggplot2 package (version 3.4.2)^83^, the ggpubr package (version 0.6.0)^84^, and the Hmisc package (version 4.5-0)^85^. Shapiro-Wilks tests, ANOVA analyses, t-tests, were conducted with the rstatix package (version 0.7.2)^86^. Robust ANOVAs and robust post-hoc tests were conducted using the WRS2 package (version 1.1-4)^87^. Correlation analyses were completed and visualized with the ggcorrplot package (version 0.1.3)^88^. Partial correlations were implemented using the ppcor package (version 1.1)^89^. Logistic regression analysis was carried out using the lmer4 package (version 1.1-33)^90^.

Before assessing for group-comparisons, data was inspected for normality of distribution using Shapiro-Wilks test. If data was normally distributed, ANOVA and t-tests were conducted, and if assumptions for normality were not met (Shapiro-Wilks p>0.05), robust ANOVA and robust post hoc tests were conducted based on 20% trimmed means and 2000 bootstrap samples. Spearman Rank tests were used for correlation analyses. Correlations were not controlled for multiple comparison due to the explorative nature of the correlations and the small sample sizes of each groups, which would otherwise lead to excessively conservative results. Correlations across all participants are presented in the supplementary material. In all the tests, a p-value<0.05 was considered significant. Potential outliers with values >1.5 times the interquartile range of the respective score were identified and excluded from the dataset. Outlier exclusion has only been applied to those analyses were stated.

## Results

### Task learning

To assess whether participants understood the task we compared task outcomes for each group contrasting the first and the last 50 trials. There was a significant effect for first and last trials, but not for group or the interaction between both (see supplements for details), indicating that all groups performed similarly independent of the strategy. Interestingly the results indicated that all participants did worse on the last 50 trials.

### Stay probability – Group differences

Stay probabilities for ideal model-free and model-based behavior are displayed in Figure 2a and actual stay probabilities in the current task are displayed in Figure 2b. The visual inspection and comparison to simulated behavior revealed that controls and OCD patients employed model-free decision-making behavior, MDD patients used a hybrid strategy and schizophrenia patients were largely random in their choice profile. The mixed factor ANOVA identified significant main effects of group (F(3, 96)=6.61, p<0.001; η_p_^2^=0.17) and reward outcome in previous trial (F(1, 96)=43.08, p<0.001; η_p_^2^=0.31), and significant interaction effects between group and reward outcome in previous trial (F(3, 96)=2.99, p=0.035; η_p_^2^=0.09), and transition type in previous trial and reward outcome in previous trial (F(1, 96)=6.46, p=0.013; η_p_^2^=0.06). See Supplementary Table 1 for post hoc tests.

**Figure 2.**
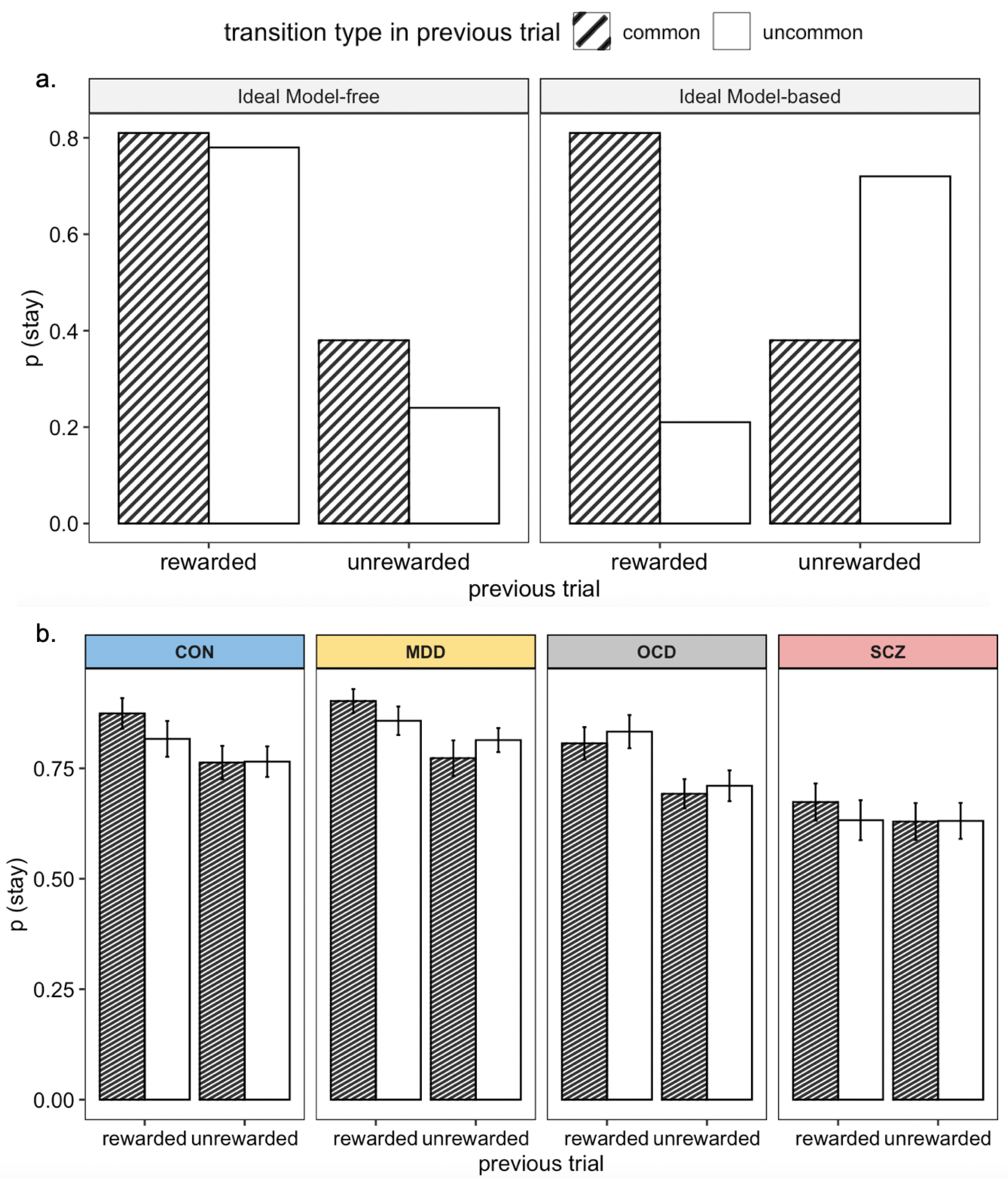
Ideal and actual stay probabilities. Note: (a) Ideal, simulated model-based decision-making behavior: Model-free reinforcement learning predicts that a first-stage choice yielding a reward is likely to be repeated on the upcoming trial, regardless of a common or an uncommon transition; Ideal model-based decision-making behavior: Model-based reinforcement learning predicts that an uncommon transition should affect the value of the next first stage option, leading to a predicted interaction between reward and transition probability; (b) Rewarded trials, independent of transition show higher stay probabilities across healthy controls, MDD and OCD suggesting increased model-free behavior; the plot slightly displays an influence of both strategies in MDD; and random choice behavior in SCZ patients.

### Logistic regression results and correlations

The robust mixed ANOVA revealed a significant effect for the decision-making strategy (F(1,50.51)=20.43, p<0.001, η_p_ ^2=^0.42), but no effect for group or an interaction. Post-hoc tests revealed that all participants favored model-free decision strategies (Mean=0.034, SD=0.08) over model-based decisions (Mean=-0.03 e^-9^, SD=0.01), psihat=0.05, p<0.001, 95% CI [0.04, 0.08]. Based on our hypotheses, we investigated group differences within model-free and model-based behavior separately comparing individual betas for reward and reward*transition, respectively. We found a significant difference for the estimate of model-free decision-making – reward beta (F(3,30.91)=3.84, p=0.03, η_p_ ^2=^0.48); but no differences for model-based behavior. Post-hoc pairwise comparisons revealed that MDD patients and OCD patients employed significantly more model-free behavior compared to schizophrenia patients (Supplementary Table 2).

Correlation analyses revealed that more model-free behavior is linked to stronger depressive symptoms (r=0.34, p=0.004, Figure 3a); and in a trend that more model-based behavior would be associated with less anhedonia (r=-0.21, p=0.075, Figure 3b). Within groups separately, we found a negative correlation between reward beta and HAMD in MDD patients (r=-0.52, p=0.02), indicating that more model-free behavior is associated with fewer depressive symptoms. This correlation is driven by two individuals, see supplements for further analysis.

**Figure 3.**
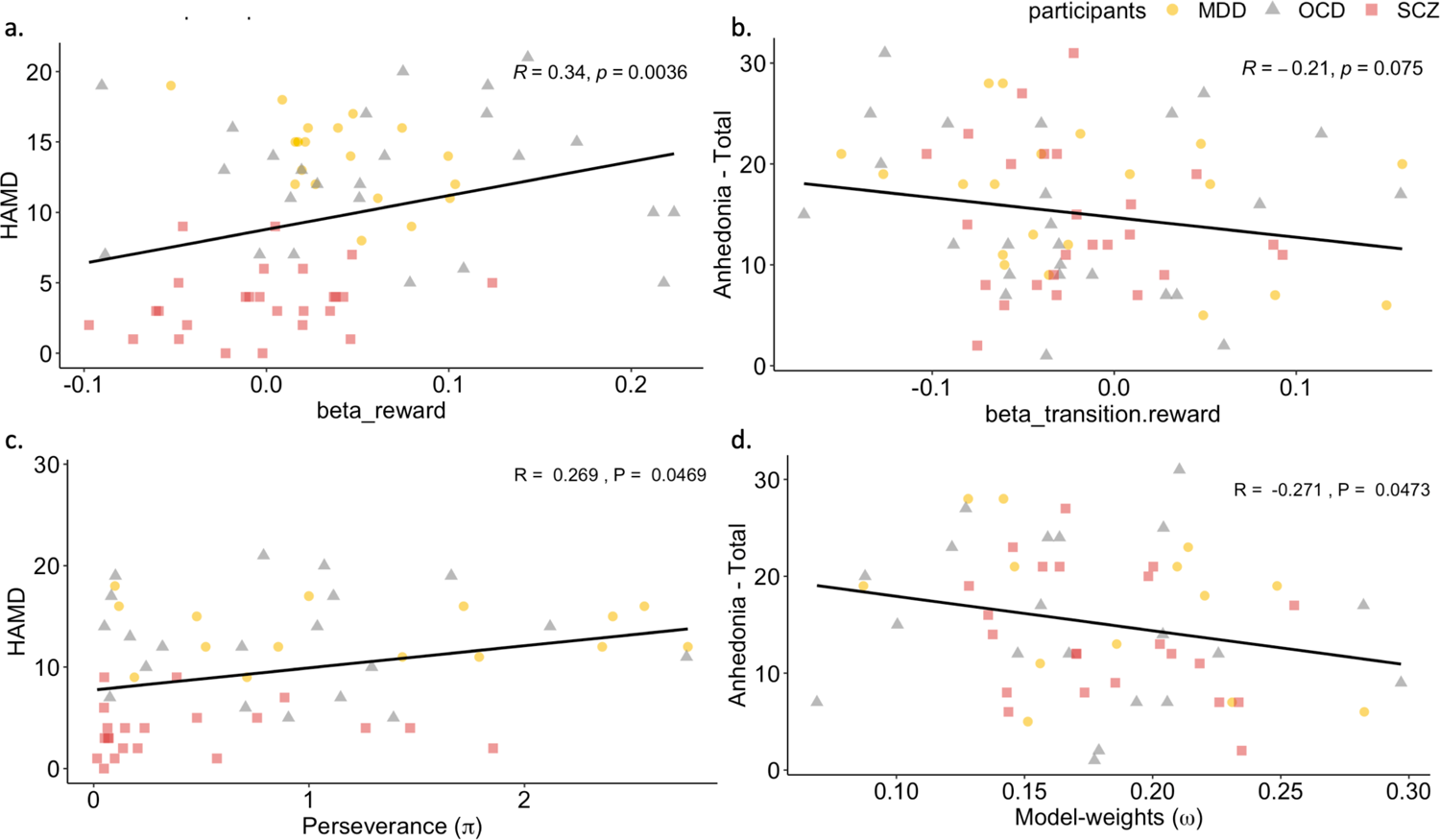
Correlations between model-free and model-based behavior and clinical scores across all groups. (a) reward beta estimate from the logistic regression and HAMD (b) reward x transition interaction beta estimate from the logistic regression and Anhedonia total (c) Perseverance (π) from the computational model and HAMD (d) Model-weights (⍵) from the computational model and Anhedonia – Total.

### Modeling parameters – Group differences

The computational modeling analysis allowed the investigation of differences in six computational parameters underlying the decision-making behavior. Significant group differences (Figure 4) were identified in the learning rate at stage one ⍺-1 (F(3,22.04)=9.12, p=0.01, η_p_^2^=0.43), but not in the learning rate at stage two, in the choice randomness at stage one β-1 (F (3,29.16)=3.52, p=0.06, η_p_^2^=0.41) and stage two β-2 (F(3,30.07)=9.68, p=0.002, η_p_^2^=0.68), and the perseverance π (F (3, 29.58)=11.05, p<0.001, η_p_^2^=0.51). Although not showing any group differences the parameter model weights (⍵) indicating model-free vs model-based behavior showed clear evidence for model-free behavior in all groups. Post-hoc analyses (Supplementary Table 3) showed that schizophrenia patients learned least at stage 1, while there were no differences at stage 2. Furthermore, they revealed that OCD patients were significantly more random at stage 1 compared to controls and MDD patients, while schizophrenia patients were more random at stage 2 compared to all other groups. Lastly, the post-hoc analyses showed that perseverance was lowest in schizophrenia patients compared to all other groups.

**Figure 4.**
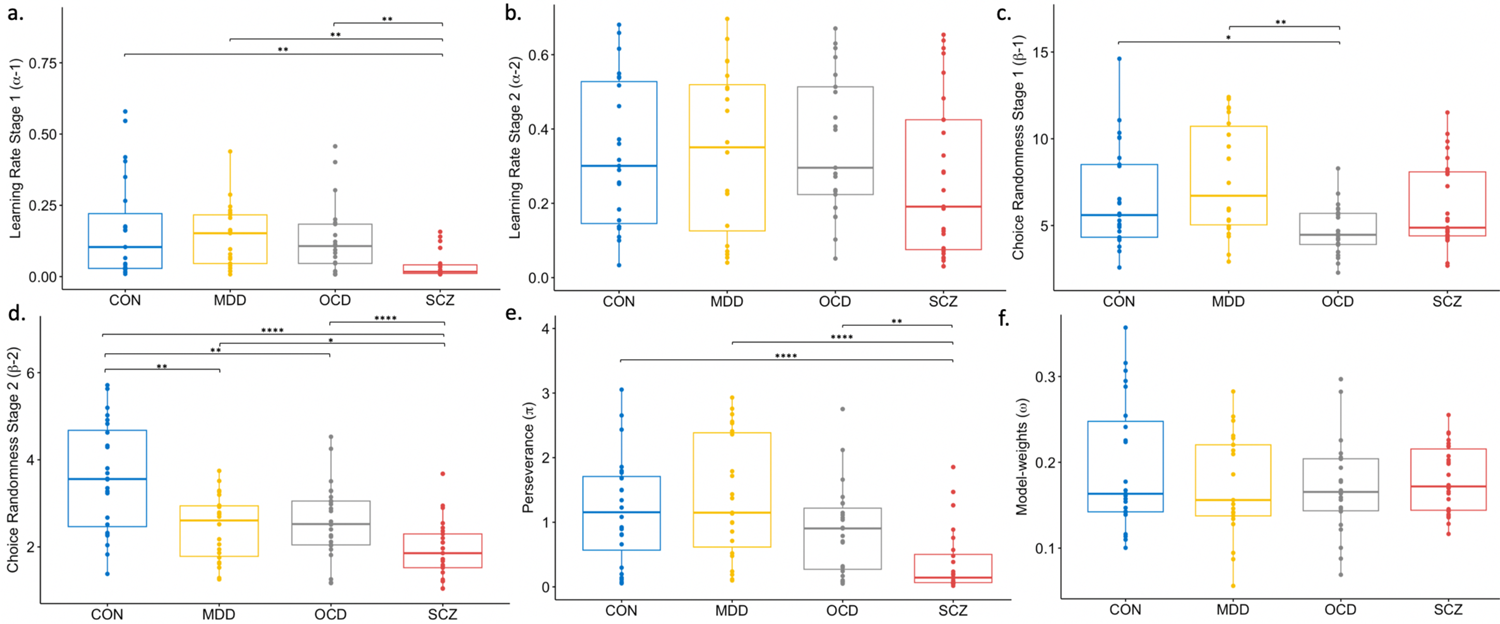
Group differences in model-parameters. Note: (a) Learning rate stage 1 (⍺-1); (b) Inverse temperature stage 2 (β-1); (c) Learning rate stage 1 (⍺-1); (d) Inverse temperature stage 2 (β-2); (e) Perseverance (π); (f) Model-weights (⍵); Group differences are calculated with robust post-hoc tests based on trimmed means using bootstrapping method *: significant differences at p<0.05; **: significant differences at p<0.01; ***: significant differences at p<0.001;; ****: significant differences at p<0.0001; p-values are adjusted using Bonferroni correction. The individual box plots represent the minimum, maximum, median, first quartile and third quartile and the outliers in the data set.

Across all patients, correlation analyses revealed that greater unwillingness to adapt the decision strategy was linked to stronger depressive symptoms (r=0.269, p=0.047, Figure 3c); and that more model-based behavior would be associated with less anhedonia (r=-0.271, p=0.047, Figure 3d). Within groups separately, we found that more model-free behavior is associated with less anhedonia (r=-0.48, p=0.038), and in a trend that more model-based behavior is also associated with less anhedonia (r= −0.39, p=0.098). Additionally, we explored association between modeling parameters and beta estimates from logistic regressions. The results are presented in the supplementary materials.

## Discussion

In this study, we investigated disorder-general and disorder-specific decision-making patterns and their associations with symptoms in individuals diagnosed with MDD, OCD, and schizophrenia using two computational approaches. We found that controls and OCD patients predominantly utilized model-free decision-making, while MDD patients exhibited a hybrid strategy, and schizophrenia patients showed largely random decision-making. These results were confirmed by regression and Bayesian analyses. Regression analysis revealed a stronger reliance on model-free strategies in OCD and MDD patients compared to schizophrenia patients. Bayesian analysis showed that schizophrenia patients had the lowest learning rate and perseveration, indicating random behavior, while OCD patients exhibited the lowest influence of reward at stage one, and schizophrenia patients showed the least influence at stage two. Importantly, both approaches revealed consistent symptom correlations across all patients showing that more model-free decision-making was associated with stronger depressive symptoms, whereas more model-based decision-making was linked to lower anhedonia. These disorder-general alterations spanning various psychiatric disorder offer insights into similarities and potential targets for interventions, especially for co-occurring symptoms.

Whereas all groups showed a preference for model-free behavior inspecting the stay-probabilities, the MDD group showed to some extent hybrid model-free decision-making behavior. This finding was quantified by the Bayesian computational parameters, which revealed that choice randomness in MDD patients was lowest compared to all other groups at the first stage, whereas choices at the second stage were more random compared to controls, but less random compared to all other groups. Although this is, to our knowledge, the first time that model-free and model-based decision-making has been investigated in patients with depression using a task that requires both strategies such as the two-step task, research from healthy individuals with depressive symptoms indicated similar results. Blanco and colleagues^91^ showed in an earlier study using a leap frog task, that depressive participants used a reflexive, model-free decision-making strategies which did not integrate environmental changes and explored randomly. Similarly, Heller and colleagues^67^, suggested that individuals with stronger depressive symptoms transitioned to model-free behavior when exposed to stress. This is interesting as research has consistently shown that individuals with stronger depression, or with MDD also reported higher levels of stress^92–94^ and depressive symptoms respectively. Importantly, this notion is supported by the symptom correlations that we found across all patients. These indicated a reduction of depressive symptoms with less model-free behavior and consistently a reduction of anhedonia with more model-based decision-making behavior. Interestingly, within the MDD group alone we found a negative correlation between model-free behavior and depressive symptom, indicating, fewer symptoms with more model-free behavior. This is in contrast to the overall correlation. We argue that while most MDD patients align with the correlation reported across all patients, only two individuals are driving this negative correlation, without whom the correlation within the MDDs is no longer significant, but remains across all patients (see supplementary materials for supportive analyses). Also, it has been shown that reward processing is sensitive to medication doses^95^ which could additionally impact the correlation.

Our results, furthermore, showed that the OCD group used stronger model-free behavior compared to controls, which is consistent with the literature^31,41,96^. For example, Voon and colleagues^41^ found that OCD patients showed less goal-directed, model-based and more habitual, model-free choices to rewarding outcomes also using the two-step task. The increased model-free behavior was linked to stronger obsessions^41^. Thus, this overreliance on habituated behavior^30^ has been linked to the inability of inhibiting established responses, especially in novel situations. This may be due to difficulties in shifting attention from one aspect of a stimulus to another, and in suppressing or reverting to previously rewarded responses^97,98^. As individuals with OCD also exhibit neurocognitive deficits in attentional and extra-dimensional set-shifting, affective set-shifting and reversal learning, as well as task shifting^99,100^, this cognitive inflexibility may underlie the inability to dynamically balancing between model-free and model-based decision-making. While all analyses in our study confirmed a clear reliance on model-free decision-making behavior, the Bayesian computational parameters also revealed that decisions made by OCD patients were largely random, as seen by the decreased impact of preceding rewards (i.e., increased choice randomness) and more choice switches (i.e., reduced perseverance). These computational parameters indicate increased random exploration, which has been linked to impulsivity^101^ and compulsivity^102^. Although we did not find that more model-free or less model-based behavior was linked to stronger obsessions and compulsions, we did find an association with stronger depressive symptoms when patients made more model-free and less model-based choices.

Confirming our hypothesis, schizophrenia patients in our study made largely random choices, which was especially apparent in rewarded common transitions. This is partially in contrast to earlier findings by Culbreth and colleagues^63^ showing that chronic schizophrenia patients were impaired in model-based behavior, but employed model-free decision-making in the same task, and to findings by Waltz and colleagues^103^, who reported that schizophrenia patients showed alterations in goal directed explorations but not random exploration, whereas more simple probabilistic learning tasks have consistently shown increased randomness in decision-making^38,55^. The Bayesian computational parameters in the present analysis clearly showed increased random exploration through decreased impact of preceding rewards (i.e., increased choice randomness) and more choice switches (i.e., reduced perseverance), which is very similar to the behavior seen in OCD patients within this study. Interestingly, it seems that the schizophrenia patients in our sample were particularly insensitive to the outcome of their choices, causing a disruption of model-free decision-making strategies. The insensitivity to reward has been shown across all stages of schizophrenia and has been associated with negative symptoms, especially anhedonia, and cognitive deficits^55,61,104,105^. Thus, our findings are in line with previous literature elucidating that these patients generally possess an impairment in reward anticipation and effort-based decision-making^8,61^. Completely random choice behavior on the two-step task might be a useful indicator for schizophrenia patients in remission of positive symptoms, however, this requires further exploration. Again, this is supported by the correlations we found across all patients, showing that more model-free and less model-based behavior is linked to more depressive symptoms and higher anhedonia.

Despite some differences across the groups, the results seem to converge in showing that higher reward effect or sensitivity, as indicated by less randomness and more model-based decision-making, are associated with a reduction of symptoms, especially anhedonia and depressive symptoms. This is in line with the previous literature suggesting impaired reward processing is associated with such symptoms across different psychiatric disorders^56,61,106–110^. This is an important finding of this transdiagnostic study, as anhedonia and depressive symptoms occur in all three disorders. The results seem to indicate a shared behavioral mechanism which underlies these symptoms, suggesting that model-based, or goal-directed decision making, is beneficial for reducing symptoms. This provides a potential starting point for a behavioral intervention in order to reduce anhedonia and depressive symptoms.

### Limitations

Whereas previous studies were able to elicit both model-based and model-free behavior with the original set up^41,59,69^, using the standard two-step decision-making paradigm, we were unable to induce model-based behavior even in healthy controls. This might be due to implicit task instructions and using Gaussian-randomized reward probabilities. Akam and colleagues^111^ and Da Silva and Hare^112^ were both able to induce model-based behavior changing reward distribution and paradigm instructions, respectively. The results of the Bayesian model elicited fewer and sometimes contradictive correlations with symptoms. One reason may be that we fitted the model under one hierarchical prior across all subjects independent of group which allows comparability between groups, which is necessary to understand distinct and shared associations. Previous work^55,113^ indicated clear differences in the parameters when using group specific vs. group general priors. The rational however for fitting the model across all participants is that this would allow us to investigate the most robust, albeit very conservative, model. None of the analyses has been controlled for medication. As we use a transdiagnostic sample, we were unable to calculate general equivalent doses combining medication for different symptoms and disorders. Furthermore, the correlations are not adjusted for multiple comparisons, given the exploratory nature of the analysis and the small sample sizes. Any significant findings will necessitate validation in subsequent studies. This study only tested one decision-making paradigm, a broader set of paradigms would further increase the significance of the findings, and allow a more fine-grained investigation of mechanisms underlying shared symptoms.

## Conclusion

Taken together, this study revealed that while all groups performed more model-free decision making, there were differences in learning rates and randomness of choices. Importantly, the results indicated that more model-free behavior was linked to an increase in depressive symptoms and more model-based behavior was associated with lower levels of anhedonia across all patients. Thus, this study highlights important general and specific decision-making alterations in individuals with MDD, OCD and schizophrenia and that co-occurring alterations and symptoms potentially share underlying behavioral mechanisms.

## Data Availability

The data for this study is available upon reasonable request to the corresponding author.

## Acknowledgements

We thank all participants for their time and dedication.

## Conflicts of interest

None of the authors reported any conflict of interest.

## Notes

### Competing Interest Statement

The authors have declared no competing interest.

### Funding Statement

This study did not receive any funding

### Author Declarations

Ethics committee of the Technical University of Munich gave ethical approval for this work.

### Summary of Updates

In this revision, the text has been shortened and generally revised. A new title has been provided and power analysis has been added.

