## Supplementary Material for "Investigating disorder-specific and transdiagnostic alterations in model-based and model-free decision-making"

1. **Task description**

All participants completed the two-step task replicated from the study by Daw and colleagues^69^ (Figure 1), which is a reinforcement learning task allowing the discrimination between model-free and model-based decision-making. All participants performed this task first outside and then inside an fMRI scanner, but only data from outside the scanner was used in this study. For imaging results see ^59^, Brandl et al. (in submission), Sen et al (in submission). The goal of the task was to maximize the monetary reward over 200 trials. Specifically, each participant completed four blocks of 50 trials each. Each trial had two decision stages. In both the stages, the participants were asked to select one of two presented fractal images. Depending on their choice in stage one, a specific set of two images of two possible sets was presented on stage two. Transitions between stage one and two were probabilistic. On common transitions, choosing image A on stage one led to ‘set a’, and image B led to ‘set b’ with probability of 70%. In 30% of the trials, an uncommon transition occurred when image A led to ‘set b’ and image B led to ‘set a’. Common and uncommon transitions were randomly distributed across all the trials. In stage two, participants again were required to select one image, which ultimately led to receiving a reward or no reward. Transition probabilities were fixed across the experiment; however, the reward probability changed slowly and independently based on Gaussian random walks within the range of 0.25 to 0.75 (Figure 1).

Based on the feedback in stage two, the participants were expected to develop a strategy that maximizes the chances of obtaining a reward. For a model-based decision, the subject must use knowledge of transition probabilities between the states to infer the likelihood that a particular action will lead to a reward. In model-free decision-making, however, the transition behavior between the states is not considered and the current value of each action is updated on the basis of whether actions were rewarded in the preceding trial. For example, when receiving a reward after an uncommon transition, a model-based agent will be expected to change her decision in the first stage on the next trial (i.e., low stay probability), as she is aware that the other option on stage one has a greater likelihood of producing the same set as the last trial. A model-free agent however will repeat the same selection on the next trial as she is solely driven by the outcome (i.e., high stay probability).

2. **Coding of stay-probabilities**

Stay-probabilities per trial starting in the second trial were coded with 1 for stay and 0 for shift. Model-free behavior would be characterized by repeating (stay: 1) the same choice on stage one at the next trial if the last trial resulted in a reward, and by changing (shift: 0) the choice on stage one at the next trial if the last trial did not result in a reward. Transition probabilities are not considered. Model-based behavior, on the other hand, would be characterized by repeating (stay: 1) the same choice on stage one at the next trial if the last trial was either a common transition and resulted in a reward or was an uncommon transition and did not result in a reward, and by changing (shift: 0) the choice on stage one at the next trial if the last trial was either a common transition and did not result in a reward or was an uncommon transition and resulted in a reward

After assigning values to each trial, we calculated the percentage of ‘1’s for model-based and model-free behavior four categories namely ‘rewarded-common’, ‘rewarded-uncommon’, ‘unrewarded-common’ and ‘unrewarded-uncommon’ for all participants and plotted histograms.

Stay probabilities were simulated for ideal model-based and model-free performances to visualize what the stay probabilities would be if all the participants behaved either in an entirely model-based or model-free, and bar-plots were made. These plots were then compared visually with bar-plots made using stay probabilities computed according to the actual performance in the task.

3. **Performances in the groups based on rewards obtained in the task.**

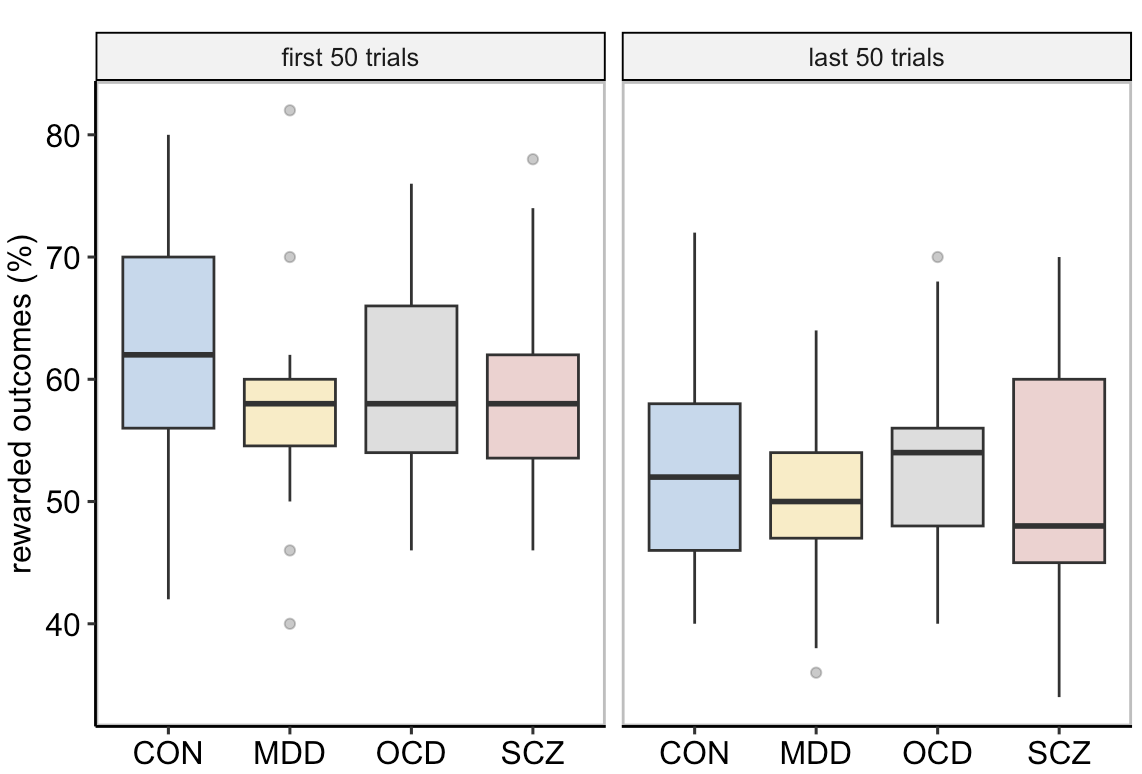

**Supplementary figure 1** **Percentage of rewarded outcomes in the first 50 and last 50 trials.**

We conducted robust mixed ANOVA based on trimmed means using the bootstrap method, with reward percentage as the dependent variable, group as the between-subject variable and pre-post trials (first 50 trials (Mean=59.5, SD=8.36, 95% CI [57.89, 61.21]), last 50 trials (Mean=52.2, SD=8.61, 95% CI [50.49, 53.9])) as the within-subject variable. We found a significant main effect of pre-post trials, F(1,53.07)=30.95, p<0.001, η_p_^2=^0.56.

4. **Tables for post-hoc tests**

| **Supplementary Table 1.** Post-hoc pairwise-comparisons for stay-probability analysis | | | |
| --- | --- | --- | --- |
| **Group 1 – Mean (SD)** | **Group 2 – Mean (SD)** | **t-value** | **P-value adjusted (Bonferroni)** |
| **Participants** | | | |
| CON: 0.81 (0.19) | MDD: 0.84 (0.16) | -1.275 | ns |
| CON: 0.81 (0.19) | OCD: 0.76 (0.19) | 1.676 | ns |
| CON: 0.81 (0.19) | SCZ: 0.64 (0.22) | 5.786 | <0.001 |
| MDD: 0.84 (0.16) | OCD: 0.76 (0.19) | 3.059 | 0.015 |
| MDD: 0.84 (0.16) | SCZ: 0.64 (0.22) | 7.277 | <0.001 |
| OCD: 0.76 (0.19) | SCZ: 0.64 (0.22) | 4.238 | <0.001 |
| **Previous_trial_reward** | | | |
| rewarded: 0.79 (0.21) | unrewarded: 0.72 (0.19) | 7.11 | <0.001 |
| **Participants x Previous_trial_reward** | | | |
| **CON** | | | |
| rewarded: 0.85 (0.19) | unrewarded: 0.76 (0.18) | 3.316 | 0.002 |
| **MDD** | | | |
| rewarded: 0.88 (0.14) | unrewarded: 0.79 (0.16) | 3.682 | <0.001 |
| **OCD** | | | |
| rewarded: 0.82 (0.18) | unrewarded: 0.7 (0.17) | 5.289 | <0.001 |
| **SCZ** |  |  |  |
| rewarded: 0.65 (0.23) | unrewarded: 0.63 (0.21) | 1.822 | ns |
| **Previous_trial_reward x Previous_trial_transition** | | | |
| **common** | | | |
| rewarded: 0.81 (0.19) | unrewarded: 0.71 (0.19) | 6.316 | <0.001 |
| **uncommon** | | | |
| rewarded: 0.78 (0.22) | unrewarded: 0.73 (0.19) | 3.739 | <0.001 |
| Note: Multiple pairwise independent t-tests conducted for the main effect of participants for stay probability and multiple pairwise paired t-tests conducted for the main effect of previous trial reward and interactions effects (participants*previous trial reward and previous trial reward*previous trial transition) for stay probability; ns=non-significant | | | |

| **Supplementary Table 2.** Post-hoc pairwise group comparisons for reward beta estimates | | | | |
| --- | --- | --- | --- | --- |
| **Group 1: Mean (SD)** | **Group 2: Mean (SD)** | **psihat** | **95% CI [LL, UL]** | **P-value adjusted (Bonferroni)** |
| CON: 0.02 (0.1) | MDD: 0.04 (0.06) | -0.01 | [-0.05, 0.03] | ns |
| CON: 0.02 (0.1) | OCD: 0.07 (0.09) | -0.03 | [-0.1, 0.02] | ns |
| CON: 0.02 (0.1) | SCZ: -0.0001 (0.05) | 0.03 | [-0.02, 0.07] | ns |
| MDD: 0.04 (0.06) | OCD: 0.07 (0.09) | -0.02 | [-0.08, 0.03] | ns |
| MDD: 0.04 (0.06) | SCZ: -0.0001 (0.05) | 0.04 | [0.002, 0.08] | 0.005 |
| OCD: 0.07 (0.09) | SCZ: -0.0001 (0.05) | 0.06 | [0.01, 0.1] | 0.003 |
| Note: Robust post-hoc tests for group comparisons of reward beta estimates based on trimmed means using bootstrapping method; ns=non-significant, CI=confidence interval, UL=upper limit, LL=lower limit | | | | |

| **Supplementary Table 3.** Post-hoc pairwise group comparisons for model parameters | | | | |
| --- | --- | --- | --- | --- |
| **Group 1: Mean (SD)** | **Group 2: Mean (SD)** | **psihat** | **95% CI [LL, UL]** | **P-value adjusted (Bonferroni)** |
| **⍺-1** |  |  |  |  |
| CON: 0.17 (0.18) | MDD: 0.14 (0.11) | -0.006 | [-0.11, 0.15] | ns |
| CON: 0.17 (0.18) | OCD: 0.13 (0.12) | 0.02 | [-0.09, 0.16] | ns |
| CON: 0.17 (0.18) | SCZ: 0.04 (0.04) | 0.09 | [0.01, 0.22] | 0.003 |
| MDD: 0.14 (0.11) | OCD: 0.13 (0.12) | 0.03 | [-0.09, 0.12] | ns |
| MDD: 0.14 (0.11) | SCZ: 0.04 (0.04) | 0.11 | [0.03, 0.18] | 0.001 |
| OCD: 0.13 (0.12) | SCZ: 0.04 (0.04) | 0.08 | [0.02, 0.17] | 0.001 |
| **β-1** |  |  |  |  |
| CON: 6.57 (2.92) | MDD: 7.65 (3.21) | -1.51 | [-4.14, 1.3] | ns |
| CON: 6.57 (2.92) | OCD: 4.71 (1.43) | 1.36 | [-0.21, 3.65] | 0.02 |
| CON: 6.57 (2.92) | SCZ: 6.11 (2.43) | 0.29 | [-1.75, 2.59] | ns |
| MDD: 7.65 (3.21) | OCD: 4.71 (1.43) | 2.87 | [0.63, 5.28] | 0.001 |
| MDD: 7.65 (3.21) | SCZ: 6.11 (2.43) | 1.79 | [-0.7, 4.45] | ns |
| OCD: 4.71 (1.43) | SCZ: 6.11 (2.43) | -1.07 | [-2.77, 0.33] | ns |
| **β-2** |  |  |  |  |
| CON: 3.59 (1.26) | MDD: 2.43 (0.75) | 1.15 | [0.26, 2.12] | 0.001 |
| CON: 3.59 (1.26) | OCD: 2.59 (0.85) | 1.002 | [0.103, 1.93] | 0.005 |
| CON: 3.59 (1.26) | SCZ: 1.93 (0.65) | 1.71 | [0.89, 2.59] | <0.001 |
| MDD: 2.43 (0.75) | OCD: 2.59 (0.85) | -0.14 | [-0.83, 0.48] | ns |
| MDD: 2.43 (0.75) | SCZ: 1.93 (0.65) | 0.56 | [0.07, 1.18] | 0.029 |
| OCD: 2.59 (0.85) | SCZ: 1.93 (0.65) | 0.71 | [0.15, 1.26] | 0.001 |
| **Π** |  |  |  |  |
| CON: 1.19 (0.84) | MDD: 1.39 (0.94) | -0.23 | [-1.12, 0.55] | ns |
| CON: 1.19 (0.84) | OCD: 0.87 (0.69) | 0.34 | [-0.28, 0.97] | ns |
| CON: 1.19 (0.84) | SCZ: 0.39 (0.51) | 0.91 | [0.32, 1.43] | <0.001 |
| MDD: 1.39 (0.94) | OCD: 0.87 (0.69) | 0.56 | [-0.21, 1.33] | ns |
| MDD: 1.39 (0.94) | SCZ: 0.39 (0.51) | 1.13 | [0.4, 1.83] | <0.001 |
| OCD: 0.87 (0.69) | SCZ: 0.39 (0.51) | 0.57 | [0.1, 0.99] | 0.003 |
| Note: Robust post-hoc tests for group comparisons of reward beta estimates based on trimmed means using the bootstrapping method; ns=non-significant; CI=confidence interval, UL=upper limit, LL=lower limit | | | | |

5. **Correlations between beta estimates from the logistic regression and the Bayesian model parameters**

The Spearman’s correlation analysis investigating the association between the betas for reward and reward*transition and the six modelling parameters revealed the following: in healthy controls we found a positive correlation between reward*transition beta and ⍵ (r = 0.63); in MDD patients we found reward*transition beta to be positively correlated with ⍵ (r = 0.6); in OCD patients we found a positive correlation between reward*transition beta and β-2 (r = 0.44). The respective heatmaps are displayed in the supplementary figure 5, 6 and 7.

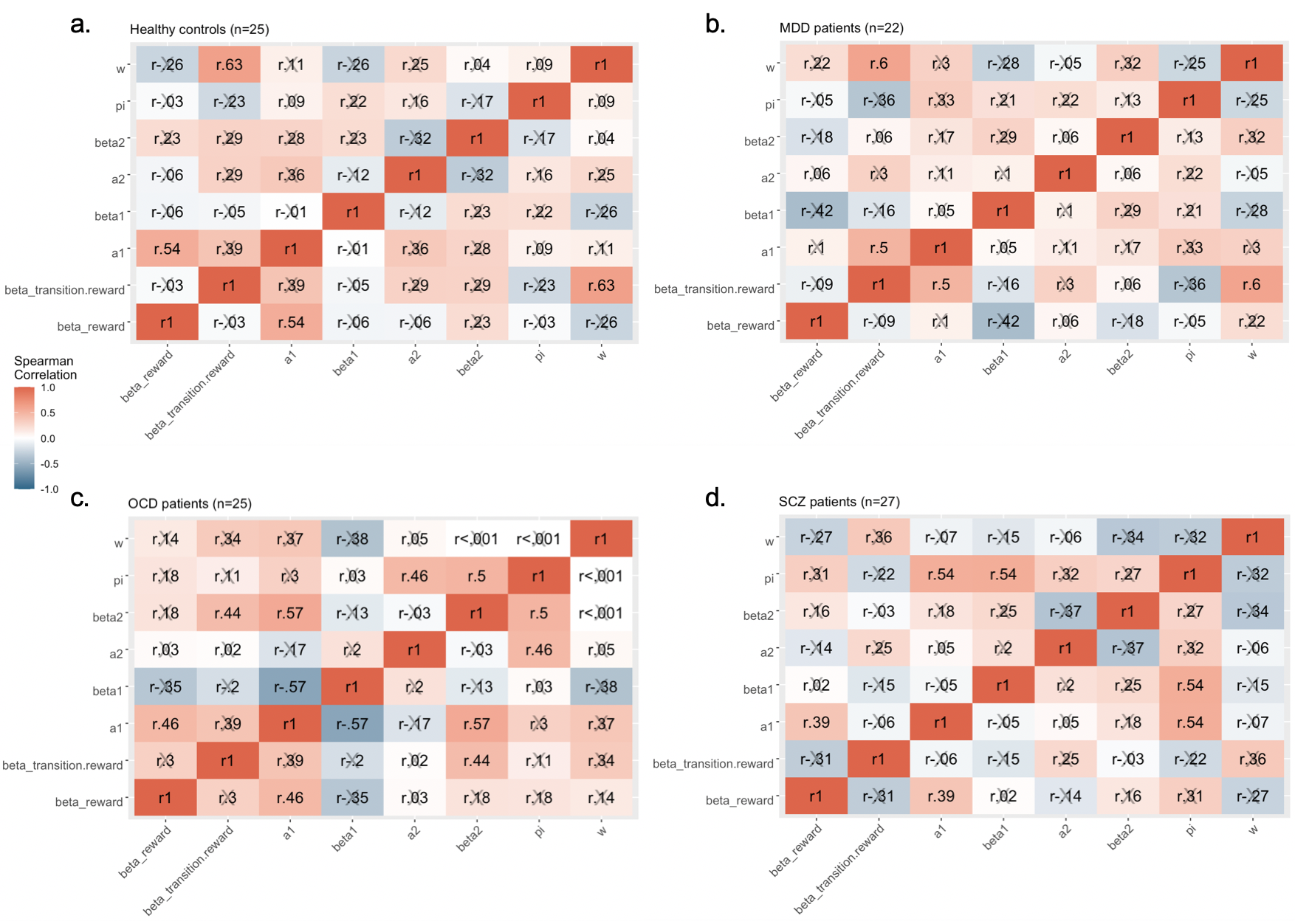

**Supplementary figure 3** **Correlations between beta estimates and model parameters for a. healthy controls, b. MDD, c. OCD, and d. SCZ patients.**

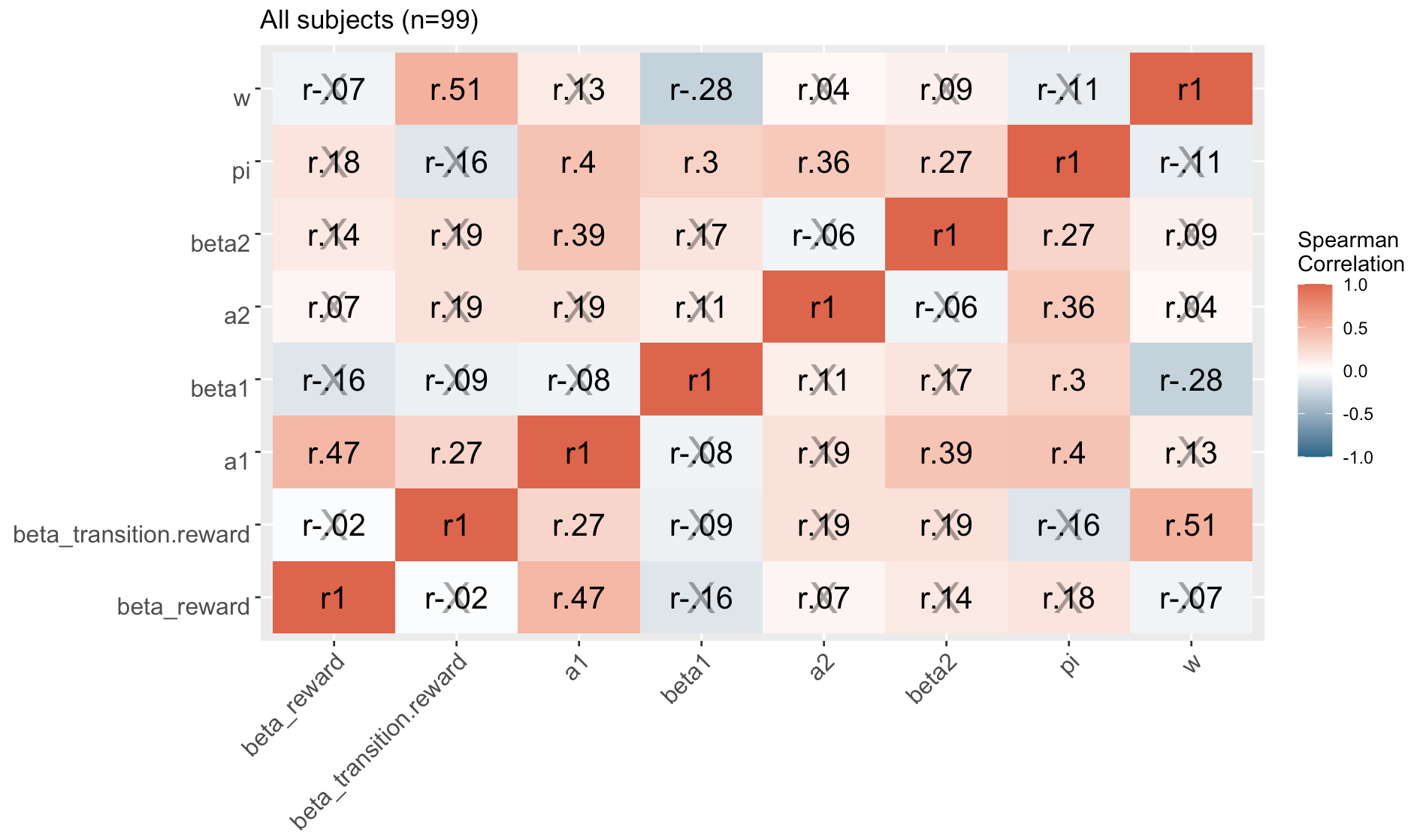

**Supplementary figure 4** **Correlations between beta estimates and model parameters across all participants.**

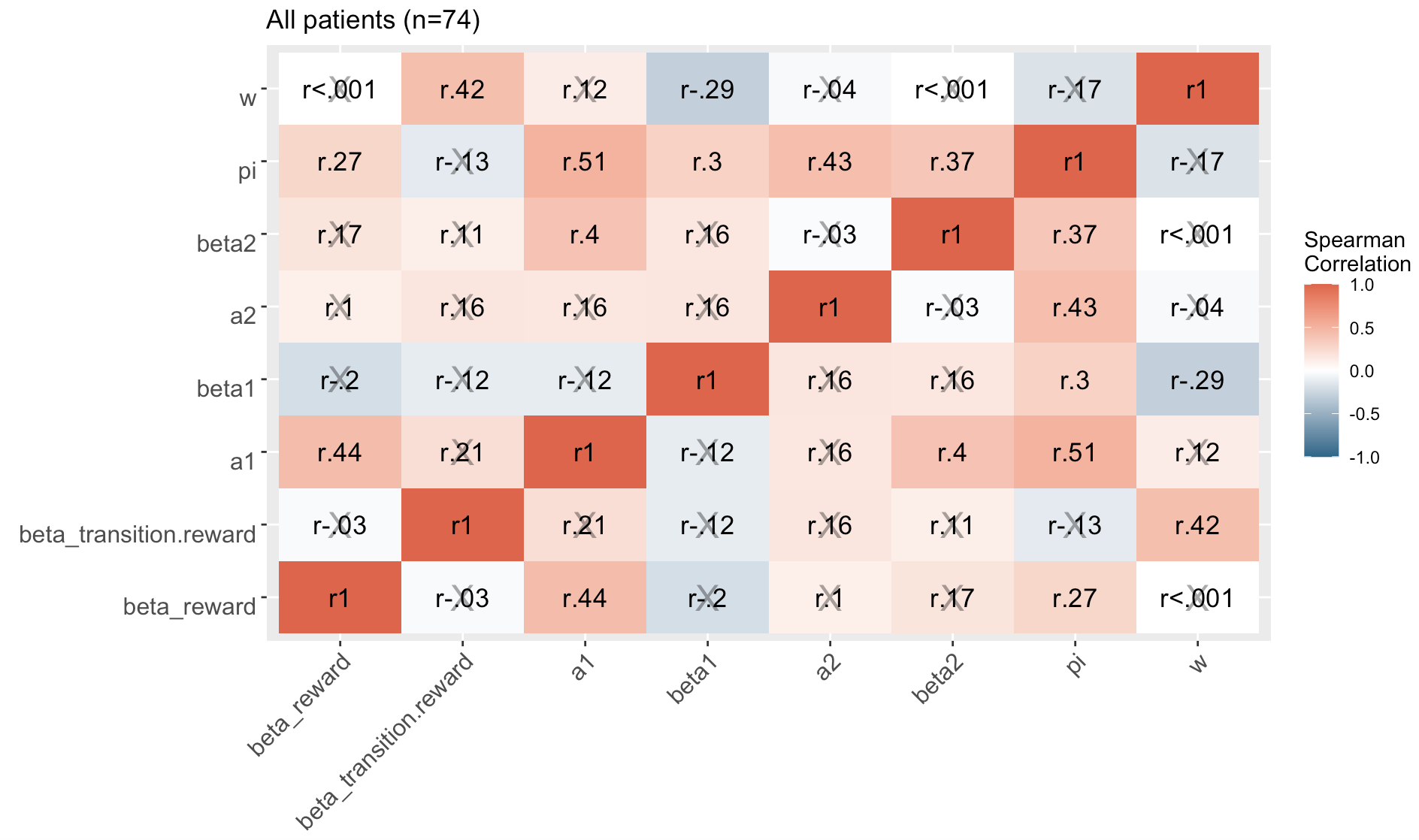

**Supplementary figure 5** **Correlations between beta estimates and model parameters across all patients.**

6. **Correlations between Bayesian model parameters and clinical scores**

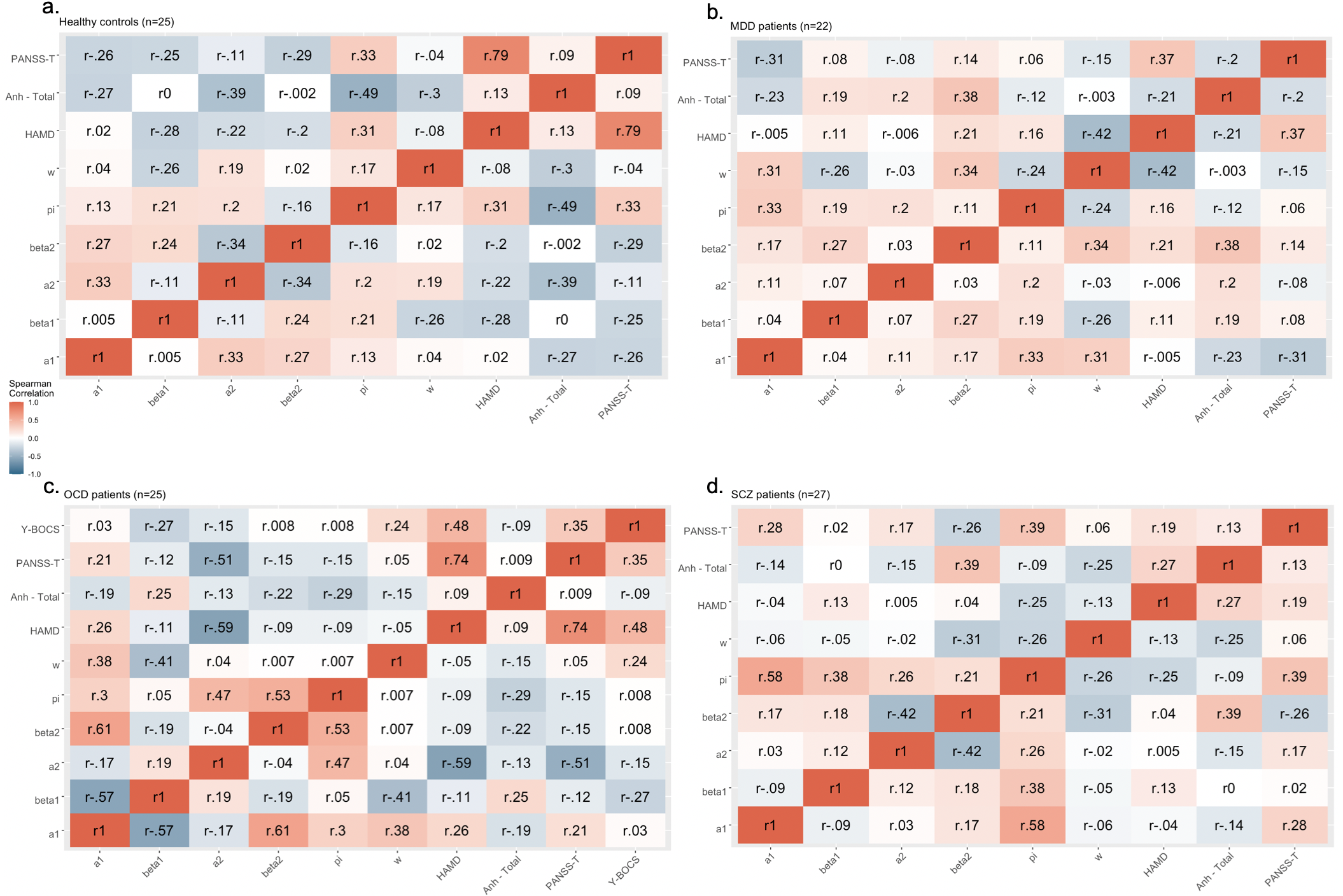

**Supplementary figure 6** **Correlations between model parameters and clinical scores for a. healthy controls, b. MDD, c. OCD, and d. SCZ patients.**

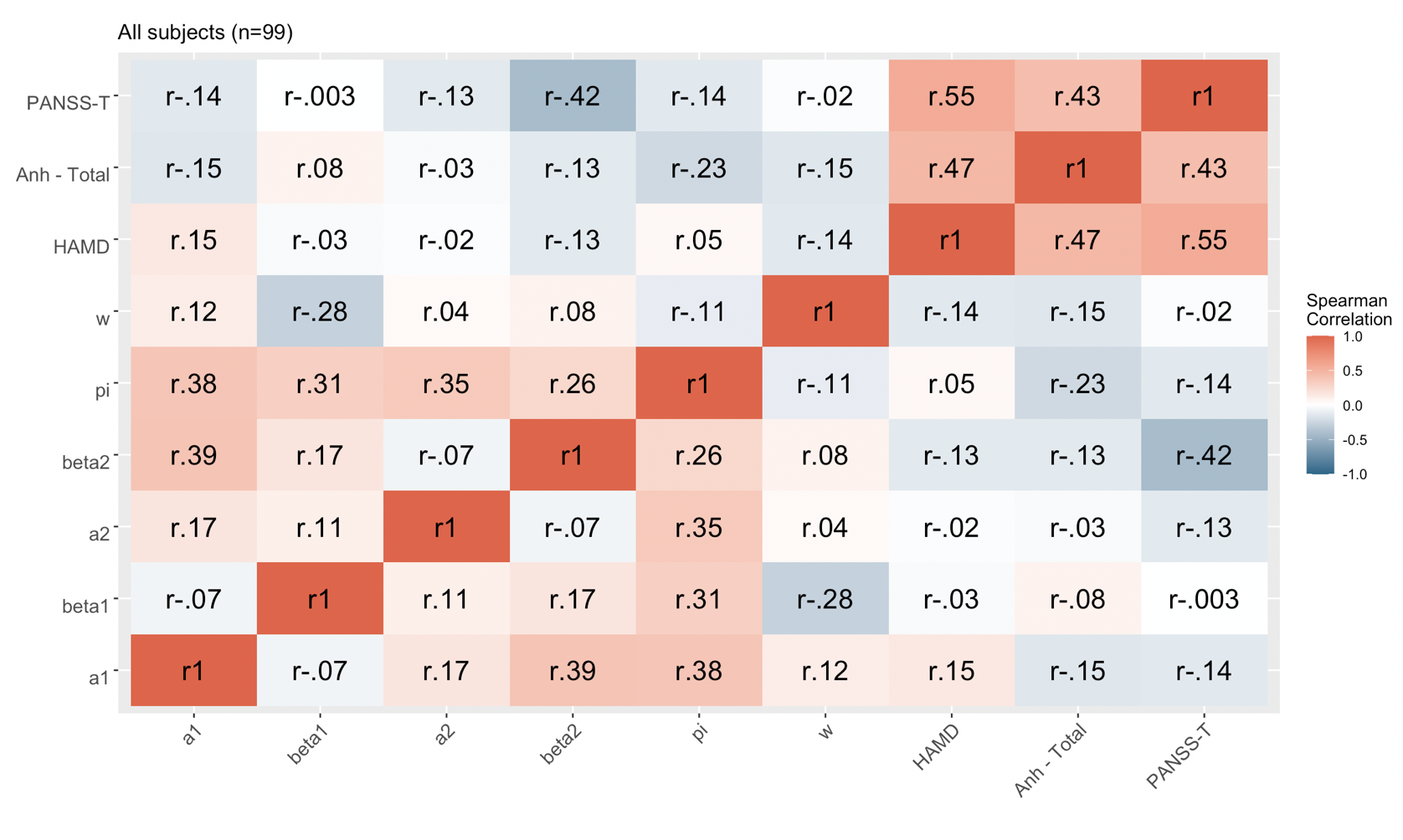

**Supplementary figure 7** **Correlations between model parameters and clinical scores across all participants.**

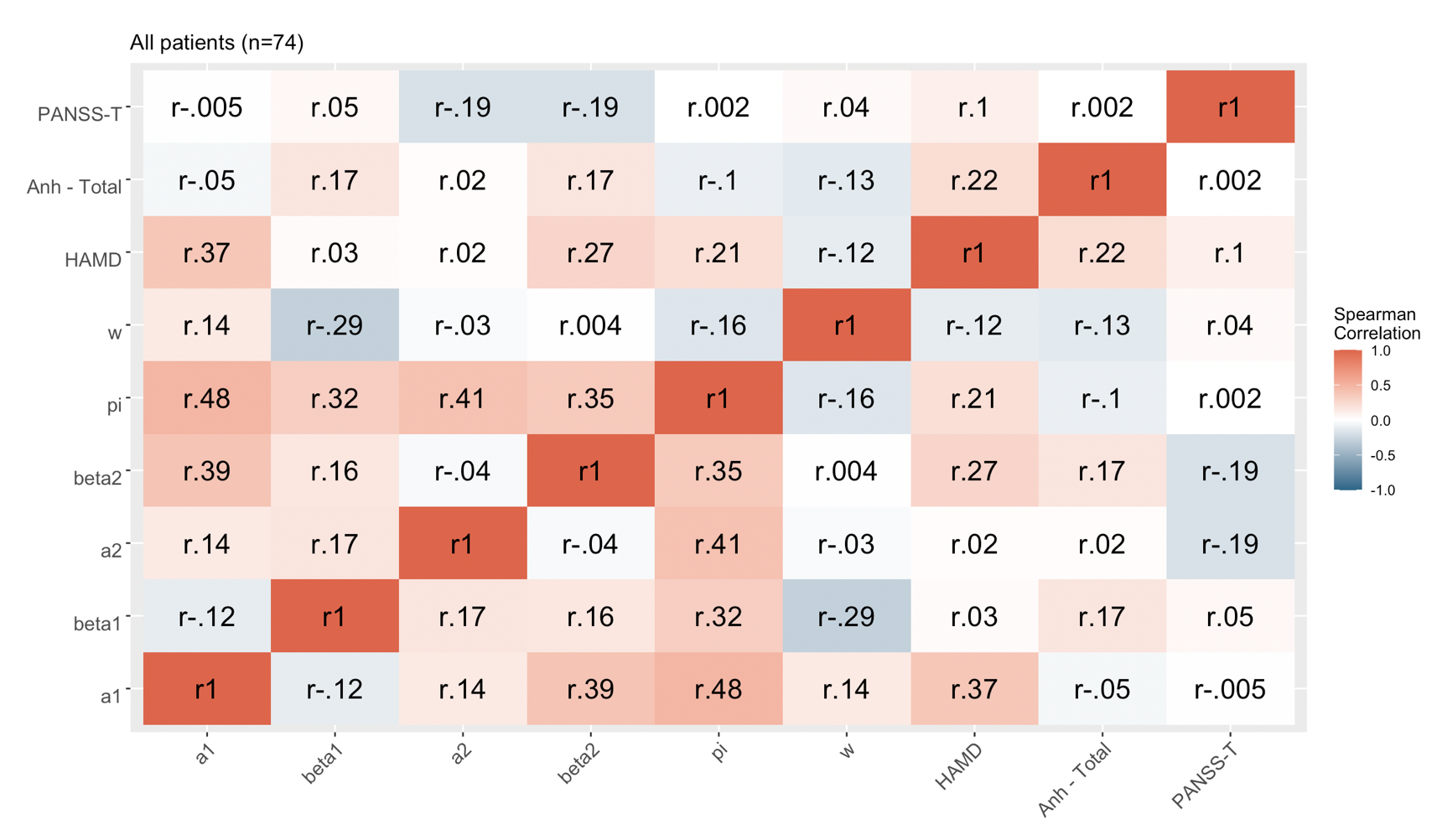

**Supplementary figure 8** **Correlations between model parameters and clinical scores across all patients.**

7. **Analysis in support of discussion**

We found a positive correlation between HAMD and reward beta estimate across all patients indicating that more model-free behavior is linked to stronger depressive symptoms in patients (r = 0.34, p = 0.004). Contrarily, within the MDD patients group we found a negative correlation between HAMD and reward beta estimate indicating that more model-free behavior is linked to reduction of depressive symptoms (r = - 0.52, p = 0.02). However, this negative association was driven by two participants who had very high HAMD scores (19 and 18) and very low reward beta estimates (- 0.05 and 0.008). After removing these two subjects, the negative correlation between HAMD and reward beta in MDD patients became less strong and was no longer significant (r = -0.33, p = 0.19) (Supplementary figure 9), whereas, the positive correlation across all patients remained significant (r = 0.41, p = < 0.001) (Supplementary figure 10).

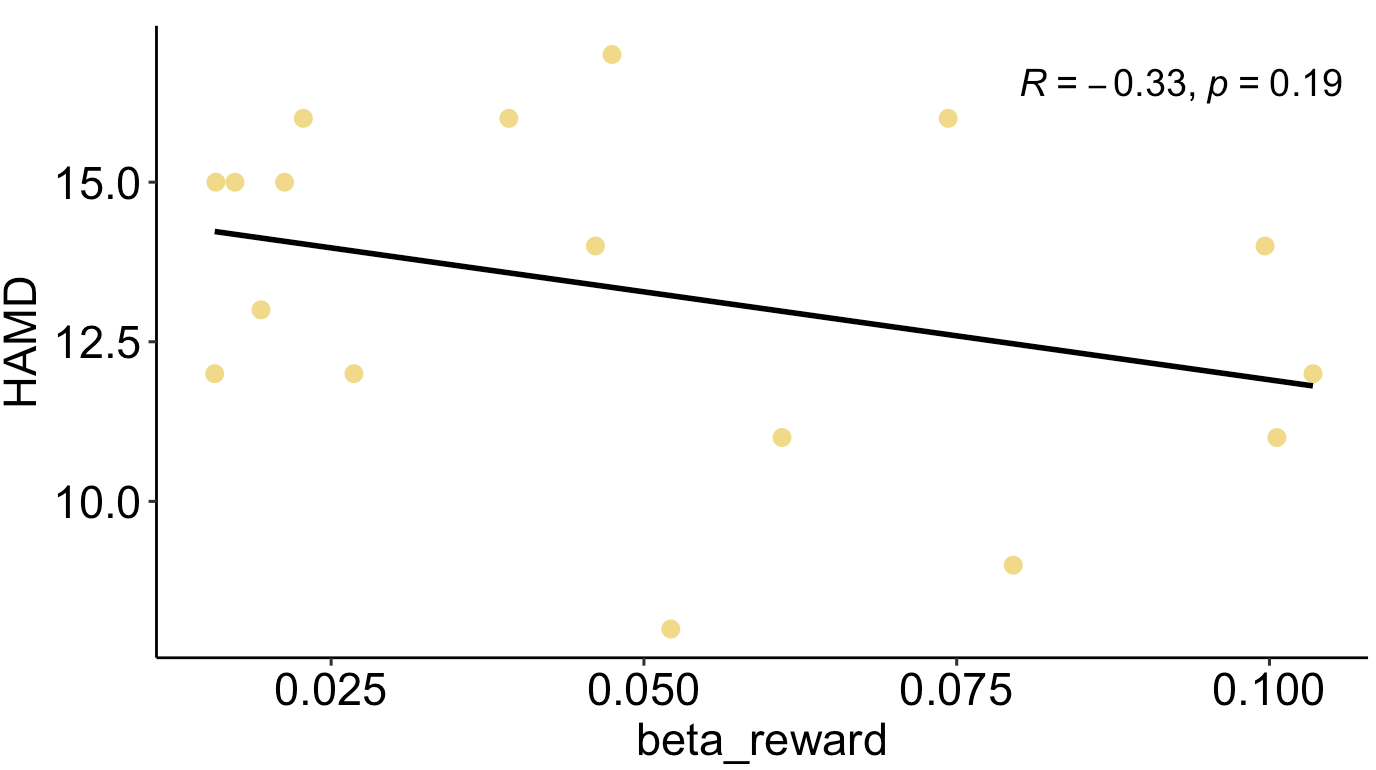

**Supplementary figure 9** **Correlations between reward beta estimate and HAMD within MDD patients after excluding two individuals with extreme HAMD and reward beta estimates.**

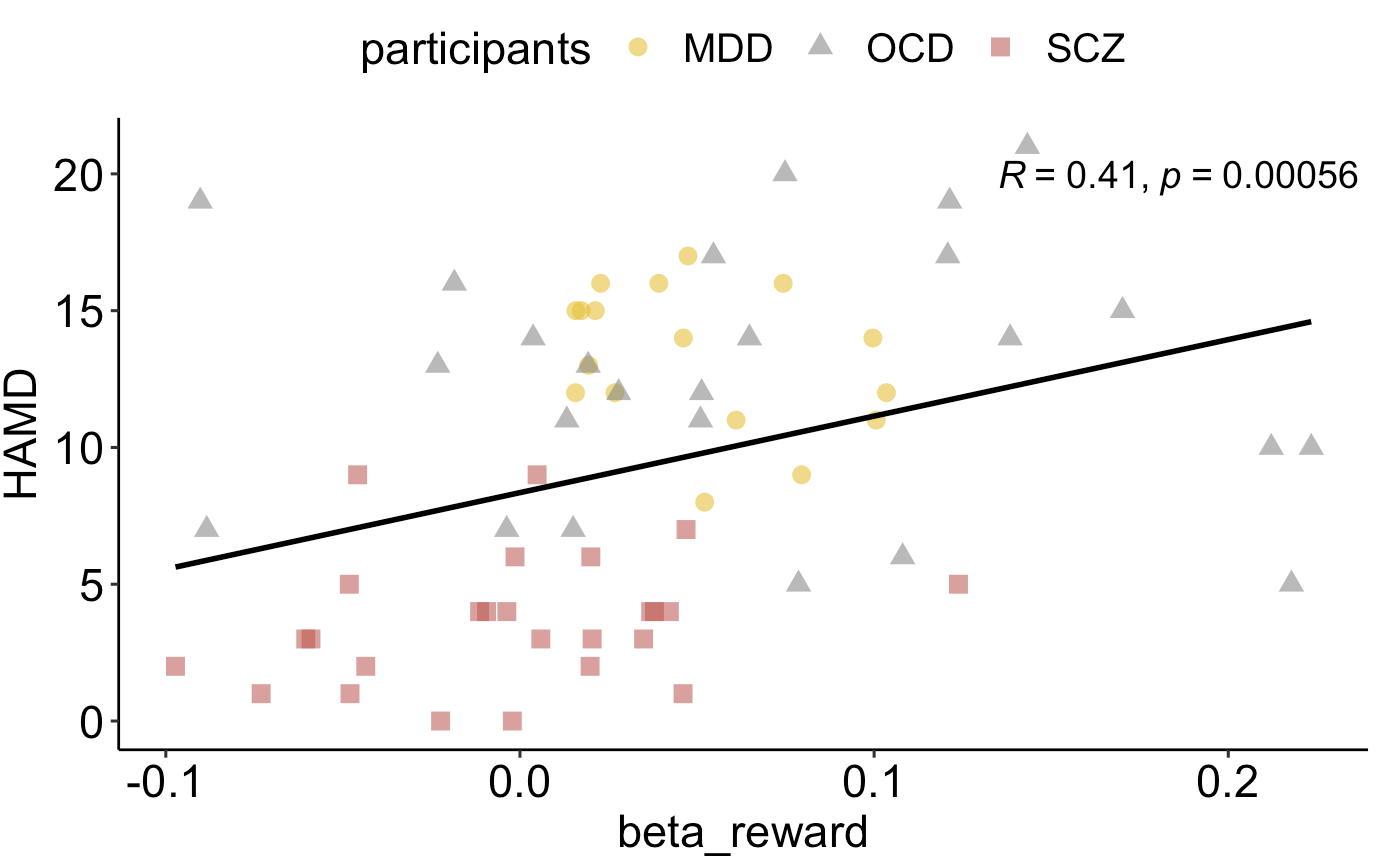

**Supplementary figure 10** **Correlations between reward beta estimate and HAMD across all patients after excluding two MDD patients with extreme HAMD and reward beta estimates.**
